# ‘Beyond Places of Safety’ – a qualitative study exploring the implementation of mental health crisis care innovations across England

**DOI:** 10.1101/2023.01.10.23284385

**Authors:** Una Foye, Rebecca Appleton, Patrick Nyikavaranda, Natasha Lyons, Ceri Dare, Chris Lynch, Karen Persaud, Nafiso Ahmed, Ruth Stuart, Merle Schlief, Xia Huong, Nick Sevdalis, Luke Sheridan-Rains, Antonio Rojas-Garcia, Martin Stefan, Jeremy Clark, Alan Simpson, Sonia Johnson, Brynmor Lloyd-Evans

**Author notes:** **Corresponding Autho**r: Dr Una Foye, Address: NIHR Mental Health Policy Research Unit, Department of Mental Health Nursing Research, Institute of Psychiatry, Psychology & Neuroscience, Kings College London, Denmark Hill, London SE5 8AF.

## Abstract

**Background:** Mental health acute and crisis care consumes a large share of mental health budgets internationally but is often experienced as unsatisfactory and difficult to access. As a result, there is an increasing move towards developing innovative community crisis services, to improve patient experience and relieve pressure on inpatient and emergency services. This study aims to understand what helps and hinders the implementation of innovative mental health crisis care projects in England.

**Methods:** Using a qualitative approach, 18 interviews were conducted with crisis care service managers exploring their experiences and views of the development and implementation of their service developed with support from an English national capital funding programme. A framework analysis was conducted informed by implementation science.

**Results:** Key facilitators to implementation of innovative crisis services included bottom-up development, service user involvement, strong collaborative working, and leadership and management buy-in. Key barriers that affected the projects implementation included the complexities of crisis care, workforce challenges and resourcing issues.

**Conclusion:** There is a recognised need to improve, update, and innovate current crisis care offers. Results from this study suggest that a range of models can help address the heterogenous needs of local populations and that such services can be successfully implemented where they utilise a whole-systems approach, involving service users and relevant professional stakeholders beyond mental health services in planning and developing the service.

## 1. Introduction

Mental health acute and crisis care consumes a large share of mental health budgets internationally but is often experienced as unsatisfactory and difficult to access. In England, a national survey conducted by the Care Quality Commission in 2021 found that over a quarter of mental health service users did not know how to access help in a crisis, and just over half did not feel they received the help they needed (*CQC, 2021*), despite a longstanding national initiative to establish crisis resolution and home treatment teams across the country *(Llyod-Evans, Paterson, Onyett, Brown, Istead, Gray, Henderson & Johnson, 2017)*. Furthermore, issues in relation to availability of care have been highlighted, with wide variations in the provision of crisis services being reported despite the nationwide availability of crisis resolution teams (*Lloyd-Evans et al., 2018*). Involuntary admissions under the Mental Health Act have risen steadily for several decades (Sheridan Rains et al), and a general rise in referrals to crisis care service was observed following the pandemic of 11% per catchment area (*CQC, 2021)*. As in many countries, inpatient and emergency are recognised as costly and often associated with poor patient experience (W*ood & Alsawy, 2016*), as well as potential harms such as loss of rights and freedoms, stigma, institutionalisation, and development of unhelpful coping strategies (J*ohnson et al., 2022; Lloyd-Evans & Johnson, 2019; Bowers et al., 2009)*. While crisis teams delivering community –based assessment and intensive home treatment continue to be core services nationally and aim to provide alternatives to admission, dissatisfaction is reported both with these teams and with choice and flexibility in the mental health care system.

Considering these challenges, improving access, quality, and choice in mental health crisis care has been the focus of several UK policy initiatives in the past decade *(HM Government, 2014; Gibson, Hamilton & James, 2016; NHS, 2019)*. Both local and national initiatives have resulted in the implementation of a range of innovative service models designed to improve quality and effectiveness of care and flexibility and integration in local crisis care systems. These have included innovations such as safe havens, crisis cafes, specialist crisis assessment services, walk in crisis centres 24-hour crisis lines, and mental health decision units (Dalton Locke). Such developments have been supported in national policy, but there are considerable gaps in evidence regarding the clinical effectiveness and cost-effectiveness of mental health crisis care, the integration of crisis care across inpatient care, post-discharge transitional care, and Community Mental Health Teams/intensive case management teams *(Paton et al., 2016)*, and innovative models have been implemented with little evidence.

A component of these national policy developments was the launch in 2018 by the Department of Health and Social Care (DHSC) in England of a call for applications for capital funding for innovative projects to improve local crisis care systems: the “Beyond Places of Safety” (BPOS) initiative *(DHSC, 2017*). Fifty projects were commissioned, which varied in scale, focus, and intended outcomes. Some were delayed due to the COVID-19 pandemic, but by late 2022, all were up and running. The models commissioned included a variety of innovative approaches to improving access, quality, flexibility, and choice. In crisis care pathways. This offers the opportunity to advance evidence on the investigate further the hanging landscape of crisis care pathways, understanding what helps and hinders innovative crisis projects in successful implementation and achieving their aims.

### 1.1. Aims

The main aim of this study is to understand what helps and hinders the implementation of innovative mental health crisis care projects in England.

## 2. Methods

### 2.1. Design

We employed an exploratory approach utilising qualitative methods. We conducted semi-structured one-to-one interviews with leads in new crisis care projects funded under the BPOS initiative project, aiming to explore their experiences and views of the development and implementation of their crisis care service.

### 2.2. Participants

The BPOS scheme funded 50 mental health crisis care projects across England, run by NHS, Local Authority, and voluntary sector providers, ranging from substantial capital-funding of new services to smaller schemes to improve or renovate existing facilities. For our study the inclusion criteria required that the funded crisis care projects had completed their BPOS funding end of year monitoring forms (a yearly form to update the funders as to the activity, achievements, and evaluation, expenditure) to ensure that the work had started and had progressed to a position where project leads were able to reflect on the implementation process. This excluded small scale projects that funded purchasing of equipment, projects that had been delayed, or projects that had withdrawn from receiving the funding. All eligible projects were invited to contribute to the study. All participants were staff members with a management and/or clinical lead role in the project.

#### 2.3. Sampling and Recruitment

A member of the evaluation team (UF) contacted the project leads for all eligible crisis care schemes to inform them about the study. Where project leads had changed, or where resource required, project teams selected the most appropriate individual to take part in the evaluation. Participants were invited to take part in an interview by contacting the lead contact (UF) who arranged an interview with a member of the research team (NL, NA, RS, RA, MS) at a time and date that suits the interviewee.

### 2.4. Measure

Topic guides were co-produced and agreed upon within study meetings with members of the MHPRU research team, experts by profession and Lived Experience Working Group (LEWG) representatives which includes people with lived experience of mental health problems, using mental health services, and/or caring for people with mental health problems. Informed by implementation science, the topic guide consisted of key questions including a) what the service was aiming to achieve and whether they think they achieved it, b) the implementation and sustainability of the project; c) the barriers that affected the implementation of the project; d) the facilitators for implementing the project; e) perceived impact and unintended consequences of the service innovation; and f) advice for other crisis care services implementing such services. In addition, we also collected brief information from participants about their work role and nature of the service they were working in, e.g., NHS Trust or third sector organisation. Participants were also asked to send the evaluation team any available reports or summary data which they were happy to share regarding changes in service use and patient flows or service outcomes, which could help the researchers understand the role of the project within its local crisis care system.

### 2.5. Procedure

We conducted semi-structured interviews online via Microsoft Teams between December 2021 and April 2022. Participants were provided with a letter of support from the crisis care scheme lead at the DHSC and were able to read an information sheet and complete an online consent form in advance of the interview. The researcher reconfirmed consent at the start of the session with recorded verbal consent taken from all participants. Interviews lasted on average 60 minutes. Audio recordings were transcribed by an external company and checked for accuracy by a member of the research team, who also anonymised all identifying information about specific people and organisations.

### 2.6. Analysis

We conducted a framework analysis (*Ritchie & Lewis, 2003*) informed by two implementation science frameworks. Our coding framework used Proctor et al.’s taxonomy of implementation outcomes (*2011)* to record projects’ evaluated or perceived acceptability, appropriateness, feasibility, adoption, penetration, implementation cost, fidelity, and sustainability. For analysis of the barriers and facilitators to implementation, the Consolidated Framework for Implementation Research (*Damschroder er al., 2009*) comprises five major domains including Intervention Characteristics, Implementation Processes, Outer Setting, Inner Setting, and Characteristics of the Individual.

Each domain consists of a number of constructs that reflect the evidence base of types of factors most likely to influence implementation of interventions (*Keith et al., 2017*). This initial stage was undertaken by seven members of the study team with each researcher charting data summaries onto the framework for each of the interviews they had conducted (UF, RA, NL, NA, RS, MS, XH). Sub-themes within each broad deductive theme from our initial framework were then derived inductively through further coding and collaborative discussion within the research team, inclusive of Lived Experience Researcher colleagues in the team (PN, CD, KP, CL).

### 2.7. Ethics

This study was reviewed by the Research Director of Noclor, the North London Research Consortium, which provides research governance services for several London NHS Trusts, who confirmed it met Health Research Authority (HRA) criteria (*2013*), to be classed as a service evaluation and formal ethical review was not required. Participants were given written information regarding the purpose of the study, aspects of confidentiality (and their limits), and the extent and limits of confidentiality including publication of anonymised quotes. Staff could decline to take part or withdraw at any stage.

## 3. Findings

### 3.1. Project Characteristics

Of the 50 projects funded through the BPOS scheme, there were 33 completed projects that met the inclusion criteria for the evaluation (60%); the remaining projects had withdrawn or had not yet completed works. Of the projects that met the inclusion criteria and were eligible to participate (n=33) a total of 18 project managers took part in an interview (55%). Of the 15 projects that did not take part this included where service managers could not be contacted, or where services had experienced high staff turnover thus staff involved in the implementation of the service were no longer available. The majority of included projects were from NHS providers (n=10) and the remaining eight were from the third sector or non-profit organisations.

### 3.2. The nature and aims of included projects

The types of projects included within the evaluation involved innovative or novel approaches such as creating or improving local crisis cafes or safe haven provision (n=7), creating online or digital places of safety such as personalised apps (n=3), and the development of multiagency hubs or pathways to improve the support offer for those in crisis (n=2). In addition, there were projects funded to improve existing spaces such as rebuilding or enhancing existing assessment suites or places of safety within mental health services. These included s136-suites which are facilities for people who are detained by the police under Section 136 of the Mental Health Act to provide a place of safety for a mental health act assessment and care (n=4), and the creation or improvement of A&E places of safety including creation of dedicated assessment suites (n=2).

Participants reported two main purposes for the projects funded through this scheme outlined by the services included within this evaluation. The first was to improve safety within acute services for individuals experiencing mental health crisis, e.g., improving s136 suites, or providing a specialist assessment suite in A&E intended to be more appropriate and safer for those in a mental health crisis. The second was to reduce use of acute services, by diverting from A&E attendance, acute admissions, or police detentions under the Mental Health Act by offering alternative, immediately accessible crisis care provision or support and safe spaces. An overarching aim of all initiatives was to improve the local crisis offer, e.g., to improve access to care or patient experience for people using services.

### 3.3. Understanding the barriers and facilitators to Implementation

Table 1 highlights the key facilitators and barriers associated to these domains that are further discussed and exemplified in the following section.

**Table 1:** Overview of Themes by CFIR Domain.

| CFIR Domain | Key themes | Facilitator or Barrier |
| --- | --- | --- |
| **Characteristics of Individuals** | Staff acceptability and belief in the service | Facilitator |
|  | Personal attributes and skills | Facilitator |
| **Characteristics of the intervention** | Bottom-up development with engagement from existing staff and managers | Facilitator |
|  | Using a flexible, person-centred, non-clinical ethos | Facilitator |
|  | Adaptability: creating a service that responds to the reality of crisis | Facilitator |
|  | Complexities of crisis | Barrier |
| **The Process of Implementation** | Planning and piloting | Facilitator |
|  | Service user involvement | Facilitator |
|  | Learning from existing services | Facilitator |
|  | Reflecting, evaluating, and executing change | Facilitator |
|  | Resources, costs, and sustainability of capital funded approaches | Barrier |
| **The Inner Setting** | Leadership and management buy-in | Facilitator |
|  | Complexities of staffing services | Barrier |
| **The Outer Setting** | Adaptability to a local context | Facilitator |
|  | Engaging key stakeholders and partners in development | Facilitator |
|  | Policy changes | Barrier |
|  | Delays due to Covid-19 | Barrier |
|  | Funding delays | Barrier |

#### 3.3.1. Characteristics of individuals

##### a. Staff acceptability and belief in the service

It was reflected that positive attitudes and buy-in from staff were important for ensuring the success to the service as these elements enhanced referrals and engagement in supporting and working in (or with) the service.

> *It was the staff group that really sort of drove the fact that they wanted to continue doing it. (Project ID 10)*
>
> *I think it’s just the passion, the dedication, the motivation, really, of staff. And it’s just going that extra mile really. (Project ID 25)*

Training staff across the existing organisation was found to facilitate this buy-in and positive staff attitudes to the new project as it allowed opportunities to build staff confidence and belief in the approach and new service being provided.

> *Staff [went] through training again on how they can sit with someone if they have the time, or how they can use it themselves.... so people engage with it. (Project ID 49)*

##### b. Personal attributes and skills

Many of the project leads interviewed who were responsible for the development and delivery of each crisis care service had experience of working on the frontline of the services they were describing. This meant they displayed knowledge and insight into the needs of the service as well as having a motivation and competence to be able to deliver meaningful change to the system. Only one participant noted challenges to staff’s acceptance of the service innovation due to the complexity of the digital approach needed to deliver it, which required external expertise.

#### 3.3.2. The characteristics of the project

##### Facilitators

###### a. Bottom-up development with engagement from existing staff and managers

The majority of projects were developed internally by members of the lead organisation, delivering them with existing staff and tailoring the national model to be adapted to the local context. This local development was felt to be a facilitator to the service succeeding as it ensured that the service was adapted to fit the context in which it was being delivered.

> *It’s quite a geographical spread, quite a rural area, so what we’ve always had to do is create two places of safety (Project ID 32)*

For services building new digital apps, it was noted that there was a need for engaging with external companies and experts in this technology due to the specialist nature of the project, however they all reported this as a collaborative approach led by internal plans and aims to ensure consistency.

Ten of the services noted service users’ involvement in the development of the project in some way. This inclusion of service users was seen to facilitate successful implementation as it further ensured that the service was meaningful to those who would be receiving care and could be shaped by awareness of the issues that previous services may have had.

> *[using a] certain level of co-production is basically, from my point of view, communities designing solutions to meet their own needs, I think, rather than us, as organisations, creating services to meet needs for people*… *we’ve had conversations about, “How can we do this better? How can we make this work?” but we are very much at the start of that journey. (Project ID 11)*

###### Creating a warm and welcoming service

Creating services that were warm and welcoming both physically and psychologically was felt by a number of services as core to their crisis care service;

> *It helps people to feel valued, and I’ve seen other places where there is, like, plastic chairs… Do you know what? If no one has taken the time to give you somewhere warm and comfortable, how do you feel welcome? How do you feel listened to? How do you feel valued? We’re just telling people that, “No matter what else is going on in your life, we’re listening to you. We value you and we want to help, if we can.” (Project ID 35)*

This was particularly important for projects in institutional settings such as A&E assessment rooms and s136 suites where people may be detained and not want to be there. Thus, having a milieu that respects the person’s needs beyond simply reducing risk was seen as key.

> *I think that no patient likes to come into a place of safety, really… all of this work is about making sure that the patient that is there is treated as a patient, not as a prisoner. (Project ID 32)*

The key aim of several assessment or s136 suite redevelopment projects was to move the crisis space outside of medical settings so that it felt more comfortable and less overwhelming. A key advantage described reported by service managers was taking away the clinical feeling associated with crisis care spaces in hospital settings such as A&E or inpatient services. This made community-based services more accessible and welcoming, making it more likely that people would turn to them for support before in their time of need, potentially seeking help before a crisis became so severe that admission seemed unavoidable:

> *It’s been received really well because if we think about the mental health and emotional well-being needs of children and young people are being heard. What they don’t want is those issues to be addressed in too much of a clinical setting. (Project ID 28)*

##### c. Adaptability: creating a service that responds to the reality of crises

An adaptable service system that can flex and bend to the needs of individuals experiencing a crisis was seen as salient by all the crisis care services. While digital places of safety were seen to provide an alternative to traditional crisis care for those in rural locations, crisis cafes and safe haven models which accept self-referral and offer out-of-hours services were considered an alternative to A&E.

Having third sector partners was perceived as a facilitator to delivering a flexible service as these colleagues seemed less influenced by restrictions and bureaucracy that could act as a barrier for people accessing services. Participants highlighted that a lack of adaptability and flexibility increases obstacles facing service users seeking help.

> *[Our lived experience colleagues who facilitate the café] tolerate individuals that […] I know clinical colleagues wouldn’t in a formal setting, “That’s not our criteria. Oh, no, we can’t have them’ … Don’t have referral criteria, as in, don’t make it harder for people to access lower-level services than it is. (Project ID 35)*

#### Barriers

##### a. Complexities of Crisis

Projects utilising digital approaches noted that there was a need to develop elements of the service externally due to the specialist nature. This created two key challenges; firstly, the external developers had limited knowledge regarding mental health and crisis care, therefore there was additional demand for the internal team to ensure a safe and sensitive tool was developed. Secondly, the need to develop externally created a conflict between what service users and staff wanted and requested, and what was logistically deliverable.

> *They might have what they think is a good idea, and it’s quite easy to verbalise, but there are more complex needs underneath that just in terms of the way they’ve engaged with the interface*… *they might be a deceptively simple thing to do but actually it is hugely complex. (Project ID 27)*

As a result, these projects were resource-heavy in both time and, cost which was felt to have a considerable impact on other aspects of the implementation process. One project noted that, as a charity, this burden was felt across the team and obliged the organisation to apply for further funding to resource their internal team to deliver the service.

#### 3.3.3. The process of implementation

##### a. Facilitators

###### a. Planning and piloting

Projects that included a pilot phase during implementation noted a number of benefits. Piloting was seen as a way to address implementation issues and plan ahead for improvements throughout the wider process.

> *We did a small sort of three-month pilot, to see if it worked. And then obviously we’d done the work to make the building better. And it’s still running now, so that’s brilliant. (Project ID 09)*

For one project using a new digital approach, piloting had an added benefit as it provided a means of build team skills and confidence in implementing the project, as well as increasing staff buy-in:

> *I think the most important thing is to start with your own team and your own staff. Get them trained. Get them piloting. We got people to act as clients and then reversed it. We had a whole morning of that, chaos ensued, but we did that because we wanted to try all the different aspects of it in terms of what could go wrong. (Project ID 41)*

###### b. Service user involvement

Ten of the projects discussed the important role of service user involvement at all stages of the service development and implementation. By having input from individuals who had experience of a mental health crisis, it was clearer to the service what could be done to create a safe and welcoming space for those in need during such periods of acute distress. This ensured that the service was a viable and useful alternative to pre-existing services.

> *Because they are a charity and we’re not employing the people with lived experience, we haven’t fallen into that trap that other places have about lack of objectivity and those issues. It’s almost like they are a critical friend, but yet people value their input and value the co-production, and everything we did as an organisation was around the co-production really. (Project ID 25)*

Community-based services such as crisis cafes were seen as being more able to involve these facilitators, while more traditional facilities such as assessment suites or s136 suites were less able as they were more restricted by elements of risk management.

###### c. Learning from existing services

Two services implementing crisis café style services found that visiting other successful services was of considerable benefit to their development of their service as this supported the developing team where guidance and evidence was limited.

> *We had [other services] come down and do some workshops with us about different models, what was good practice, what was out there. We went and visited a number of services*… *So based on that, what we came up with in the pathway was that we developed this service (Project ID 25)*

###### d. Reflecting, evaluating and executing change

Eight of the interviewees reported they were involved in a formal evaluation, and all services that received funding were required to complete monitoring forms as a form of evaluation. Other forms of feedback were used across the projects including the use of compliments and complaints, service user engagement, and informal feedback processes.

> *We have taken on board compliments and complaints, and we have worked alongside our liaison psychiatry colleagues, and dementia care colleagues, to make sure that we are as minded as possible of the recommendations around planning spaces for people living with mental health problems, or dementia. (Project ID 48)*

Many of the services noted the importance of ensuring that part of their process included capturing this feedback as part of a cyclical and iterative approach (often called ‘continuous improvement’) to obtaining insights through data and patient feedback and using this learning to guide and improve the service.

##### Barriers

###### a. Resources, costs and sustainability of capital funded approaches

The majority of projects reported that they delivered within budget. The funding was only provided for the purpose of capital costs, thus required additional revenue funding for aspects of the service such as staffing. For most services this revenue funding was sustained and sustainable within the wider services, however two of the projects noted challenges in securing these additional resources that were required to deliver a fully functioning service.

> *I think some of the difficulty was that the budget was capital funds. That limits how you’re able to use that money*… *if you set up a service then you always get that recurring cost. It would be really beneficial to have that identified ongoing resource to help with that earlier on. (Project ID 1)*

Challenges related to high staff turnover, changes in staffing following COVID-19 and changes in the scope of the project designed and that which was delivered accounted for these resourcing issues and highlighted the dynamic nature of crisis services.

#### 3.3.4. The inner setting, which includes the organizational structure, culture and climate

##### Facilitators

###### a. Leadership and management buy-in

A key facilitator for these projects was having committed and influential leadership supporting innovation and change, thus building buy-in from senior managers and commissioners. Having senior teams who were motivated to make the service work built confidence in the project among staff and made change feel achievable.

> *We had good support from our commissioners, from the managers, in terms of agreeing the bid*… *We had support within the trust and with the mental health commissioners. (Project ID 48)*
>
> *I had a new service director that came in, and we sat down, and we said, “Right, how are we going to deal with this?” So, we said, “Okay, let’s look at who we’ve got. This person has got a strength in this area*… *let’s flatten the hierarchy, and let’s unlock people’s creativity and stuff.” (Project ID 25)*

Such engagement was beneficial particularly where commissioners were engaged at an early stage in planning.

> *You know, she was involved from us drawing up the plans to make the changes to the building, she was involved at that stage, really. So that worked really, really well. (Project ID 09)*

This was felt not only to strengthen development and implementation processes, but also to support the sustainability of the services. There was belief and motivation to continue the service, thus further funding was secured to continue the service.

> *We’ve got a suicide prevention team and they actually identified that they would cough up for 18 months, to have it open on Saturdays and Sundays [because of the demand]. (Project ID 35)*
>
> *The current contract runs until the end of March, but there’s no intention from the commissioners of not continuing that beyond April. And the same commissioner has just extended another contract, that runs alongside this for another two years. (Project ID 1)*

##### Barriers

###### a. Complexities of staffing services

In the creation of new person-centred approaches for individuals experiencing mental health crisis, services set in non-mental health specialist areas, e.g., general hospital A&E settings, found that the complexity of providing such a service was a barrier to delivery. This barrier was related to the limited training that medical staff had in providing this style of crisis care.

> *The biggest thing I would say is staffing and trying to make sure that you have got appropriately trained staff to work within the environment. It is staff who are highly trained [to deliver mental health care], not just registered practitioners (Project ID 48)*

Within NHS settings in particular, staff turnover was a concern for project leads as changes in staff who championed new services could have an impact on the overall drive and motivation for the project to succeed. Exemplifying this, one service was unable to take part in the study due to high staff turnover in the service that meant no one who had been involved in the funded project was still working in the organisation.

Changes in staff were felt to impact staff motivation and drive, as well as having influence on the implementation by creating setbacks at a systemic level.

> *Every now and again, there is a change in personnel. If it’s a change in personnel in leadership, then you’ve got to rebuild all those relationships again. (Project ID 28)*

This issue of staff turnover had an impact not only at the staffing level of service but also at an organisational level in which the need for consistent leadership was central, thus any changes in this area had deep impacts that were felt through service delivery.

> *There were very organisational issues for us just in terms of leadership. Our chief executive left just at the end of March for personal reasons, but that hadn’t been anticipated, and there were issues that led up to it. That had a really big impact on us. (Project ID 17)*

#### 3.3.5. The outer setting, which comprises the economic, political, and social context

##### Facilitators

###### a. Adaptability to a local context

A number of project leads noted the importance of keeping in mind the wider context in which their service sits was necessary to ensure that they could meet the needs of the local population. This consideration was a key component in the decisions made by the three organisations implementing digital approaches, as previous projects had found that the wider rural setting made travelling to a physical location a barrier for some service users.

> *So again, the barriers for somebody in a small village 30 miles away to talk to somebody face-to-face can completely be overcome by this sort of thing [digital approaches], “You can see us face-to-face. We would love to see you and talk to you face-to-face and this is how we can do it, and it is as easy as this*… *we can do a test call, we can show you, walk you through it.” (Project ID 41)*

By understanding the wider context and needs of the local population, they were able to develop a service that would have an impact for those in need and when current alternatives such as driving to the local A&E or taking public transport to a crisis café would be challenging. One project noticed the lack of uptake in the service where evidence-based approaches were used but assumptions about the local context were made, and the local context was not accounted for.

> *My main advice would be really think about the times of opening*… *I could be open all night and I wouldn’t see a soul, ever. Because it’s just very different. So, think about your geographic demographics in terms of things like that, don’t always assume that people want to access that kind of support through nights and at weekends because actually when you look at when people attend here, it isn’t after 9 at night. The reality is, unless you’re in a city centre, which we are not, people don’t want to come out, because public transport gets worse and it’s dark. And they just don’t like that level of travel at night in more rural areas. (Project ID 1)*

As a result, this service adapted its model to address the local needs and developed satellite sites to accommodate the needs of service users in more rural areas.

> *“*…*We are really rural*… *so, we’re going to do pop-ups in the local communities, to try and take the support to them, rather than them having to come to us.” (Project ID 1)*

###### b. Engaging key stakeholders and partners in development

Services noted that having external collaborations such as police and ambulance services with key stakeholders was important for troubleshooting how to create a collaborative service integrating all key stakeholders who all have their own requirements and skill sets. This was more common in NHS services assessment and s136 suite developments where such multidisciplinary teams are required.

> *We’ve built a very clear observation area that has a nice observation window and desk spaces for three people so that the AMHPS, doctors and nurses can get in there and write their reports and have their discussions*… *and the police came and worked with us on it. It was designed to have a dual purpose; so that it was somewhere that was safe for us to manage someone’s aggressive behaviour, but it was important it was also somewhere that was welcoming. (Project ID 32)*

Not only was this networking and strategic partnering to create buy-in seen as important, but it was also felt to be a key facilitator from a care perspective. Where services were networked well within the wider crisis system there was opportunity to signpost and refer across systems thus creating opportunities to allow service user choice as well as genuine alternatives for people in crisis.

> *It’s a small community, so we know most of the practitioners that are based up in A&E, and we know the staff at the crisis team. They’re very good at being able to redirect people down to the service. And I think, when the crisis team are involved with individuals in the community, if they don’t meet their threshold, if you like, for their sorts of interventions, then again, they will signpost them down to us (Project ID 1)*

##### 5.2. Barriers

###### a. Policy changes

Overall, all services who reported detail on the adoption and penetration of their crisis care project expressed positive views regarding how the new service or changes to an existing service had integrated to their existing services and crisis offer. While the majority noted that the set-up and delivery of their project had good fidelity to their original plan and that the changes were implemented as originally prescribed, one service found that, due to policy and guideline changes in relation to Psychiatric Decision Units, the original project was no longer feasible. Therefore, the outcome was delivered differently than intended, with an urgent care hub created instead of a psychiatric decision unit:

> *Information came from the CQC, RCPsych, and from the National NHS England team around a shift away from recommending psychiatric decision units, the type that we had put into the original bid. So, we went about reviewing a slightly more therapeutic space that was attached to our Health Based Place of Safety to respond to that national directive. (ID 09, NHS Service)*

###### b. Delays due to Covid-19

As the funding was provided in 2019/20 there were considerable barriers facing all projects as a result of the COVID-19 pandemic. Some services noted delays in progress due to internal changes in staffing and ward spaces, while others noted external delays from companies doing construction work.

> *There was a construction delay [due to covid-19] some people have moved on during that time. (Project ID 4)*

In addition, services noted that demand for crisis care had increased for some of them during the COVID-19 period, reflecting the impact of the pandemic on service users’ needs and service delivery. For one A&E assessment suite, the increased demand for inpatient admissions and staffing issues in the hospital meant that service users were waiting longer in the assessment suite.

Another service providing an out-of-hours crisis café noted that during the pandemic the café was unusually busy. This led to the facilitators being overwhelmed and required problem solving regarding how to turn people away when demand was high, particularly whilst trying to sustain social distancing. While acknowledged as a challenge, they felt that this increasing demand was also a positive reflection of the service in that it was a genuine alternative to people feeling increasingly worse or having to attend A&E.

> *We only had enough money to start with to run it for three days a week, and demand is growing*… *Do we stop people coming? We can’t have more than six people visiting at a time so we’ve had to say, “If you are a regular, and actually, you’re here for a game of cards, and someone new presents, you will be asked to move on, and you understand that?” It’s not easy, but we are trying to manage it best we can. (Project ID 35)*

### 3.4. Perceived Impact

Overall, the project leads reported positive outcomes from the implementation of their project. Participants reported a range of perceived positive impacts not only on their own service but also across the crisis care systems. Table 2 below outlines the perceived outcomes according to Proctor et al.’s taxonomy of implementation outcomes (*2011)*. Outcomes presented in table 2 reflect perceptions of service leads rather than validated evaluation data.

**Table 2:** Perceived Impact categorised by Proctor et al.’s (2011) taxonomy of Implementation Outcomes.

| **Taxonomy** | **Findings** | **Example quotes** |
| --- | --- | --- |
| Acceptability | - High rates of reported acceptability for staff, with positive feedback reported across sites<br>- One site reported having improved spaces resulted in staff feeling safer at work, another reported a more positive working environment<br>- One service noted staff reported lower acceptability due to the complexity of the digital approach needed to deliver it which required outside staff to provide expertise to sustain the service<br>- Generally, staff perceived high acceptability for service users, based on anecdotal reports | "Having their own building and control over staff brought in has offered teams more control, stability and created a sense of ownership over their workplace - teams no longer have to worry about being moved on."<br>(Project ID 1) |
| Adoption | - Increased uptake of services offered reported by several sites<br>- One site reported regular attendance from staff from an outside organisation who can support service users with housing/benefits etc<br>- A few sites reported engaging well over the target number of service | "High uptake from service users - regularly 10-15 people attending each night." (Project ID 35) |
|  | users, indicating successful uptake |  |
| Appropriateness | - Staff reported positive views of working for an “innovative” service<br>- Allowing for a more collaborative approach to patient care and to look at service users’ holistic needs<br>- More appropriate setting for someone in crisis than A&E, or being picked up by the police<br>- One service noted challenges to the acceptability for service users noting that from their experience some diagnoses may make the use of a digital place of safety as less appropriate | “Seen as an asset to the crisis teams offer, known and referred to by outside partners e.g. police” *(Project ID 35)* |
| Feasibility | - Better environment, nicer building, more accessible<br>- Able to offer out of hours support: one service reported being able to provide online out of hours support to help reach people who could not access other support, or who lived far away.<br>- Allowing care to be more integrated across different services | “This allowed us- and continues to allow us, to provide that support in a way that gave more equality of access, and again, safety of discussions, safety of conversations without people needing to physically come down to the building.” *(Project ID 41)* |
| Fidelity | - Overall, the majority of services reported that the set up and delivery of the project had good fidelity with their original plan<br>- One service was an exception, reporting that due to policy and guideline changes in relation to Psychiatric Decision Units meant the original project was no longer feasible | “All aims achieved and fidelity to the funded proposal apart from receiving accreditation.” *(Project ID 09)*<br>“Information came from the CQC, RCPsych, and from the National NHS England team around a shift away from recommending psychiatric decision units, the type that we had put into the original bid.” *(Project ID 09)* |
| Implementation Cost | - The majority of services reported the implementation being within budget<br>- For the two sites which came in over budget, the extra cost was met by the | “...we couldn’t, as a charity, have invested £40,000 in that building, to have done the work to |
|  | NHS Trust<br>- A further two sites reported being within budget but had some limitations e.g. recurring costs not being covered | enable us to do this. So, to be fair, yes, we probably wouldn't be where we are now, without that funding..." (*Project ID 42*) |
| Penetration | - Several sites reported high penetration, with new initiatives integrating well into the existing care pathway<br>- In some cases, this led to decreased pressures and demand for other services<br>- Some services mentioned wanting to increase the roll out further, for example bringing it to remote areas via 'pop-up' centres | "Good usage, integrates across the pathways to relieve pressures across the directorate" (*Project ID 25*) |
| Sustainability | - All sites reported that the service would be sustained in the future<br>- In some cases, adjustments to the existing model were being planned for the future service<br>- All but one site reported that further funding had been secured to allow for the continuation of the service | "Increased demand allowed investment to fund further nights and sustain for 3 years as they saw the value" (*Project ID 35*) |

## 4. Discussion

### 4.1. Main findings

In this study, we aimed to explore what helps and hinders the implementation of innovative mental health crisis care projects in England. When taking stock of the current picture of crisis care models and evidence, Johnson and colleagues (*2022)* highlight the need for a greater understanding of how best to implement change and innovation in crisis care across a wide range of contexts. This study provides insight into the facilitators and barriers of such expansion and new models of crisis care. The use of the CFIR constructs provides a nuanced insight into the integral role of developing services with leadership, collaborative working, and staff resources at the core of the service.

Furthermore, this study notes that the need for localised, flexible, and person-centred approaches to work as effective crisis care models reflect that overall accessibility, support, and the extent to which an integrated and flexible crisis response from helpful and empathic staff is highly valued by service users (*Groot et al., 2019*). The results from this study indicate the importance of lived experience input within the development of projects as well as in their delivery with services noting that co-production and co-delivery of innovative services such as crisis cafes was central to their acceptability and sustainability among service users. Utilising such bottom-up approaches in this field is important as this ensures that the service users’ needs remain integral throughout implementation and beyond into the ongoing delivery of care (*Baker, 2001*).

While the findings in this paper suggest the strength of utilising a bottom-up approach to developing crisis care services to allow for personalisation and flexibility to account for service context, this creates a tension with the need to utilise evidence-based models. Previous literature has noted that having high fidelity to a model can be associated with better outcomes (*Llyod-Evans et al., 2016*). As reflected within this paper, the complexity of crisis means that evaluation is hard to do in such contexts, however such tensions between the local evolution of services and need for evidence-based models with well-developed methods for improving and monitoring fidelity may be helped by utilising Quality Improvement (QI) methods (*Taylor, et al., 2014*). Namely, the use of learning from existing services, piloting, and reflecting to ensure iterative change to improve services were noted as important elements that facilitated the process of implementing changes to existing services or for the creation of new services. These findings outline that it is helpful where possible to inform service development and justify continued funding through utilising this QI approaches within service improvement and innovation in mental health care.

While most of the implementation domains highlighted key facilitators to support such service change or implementation, there were clear barriers that impacted on the projects, especially the complexity of delivering crisis care in context of resource constraints which exacerbate the workforce challenges facing our healthcare systems. Most of the perceived barriers identified in this study were within the ‘Outer Setting’ domain due to these external economic, political, and social agents, which are likely to be common in any organisation as key barriers within implementation of services. Considering these challenges, the adaptation of responsive and flexible bottom-up approaches informed by service user needs and the acknowledging the nature of crisis was necessary for these projects to achieve their successes, an approach recognised as important in the delivery of complex healthcare services *(Braithwaite, 2018; Plsek & Greenhalgh, 2001)*.

Within the present study, services experiencing such resourcing issues and the lack of continuity of staff created barriers to implementation as this created a challenge in keeping momentum for innovation and changes. This likely reflects the wider context, as mental health services are experiencing workforce shortage with staff turnover being a growing issue (*BMA, 2020*). Alongside the challenges noted in the study in relation to barriers created by the COVID-19 pandemic, this adds further emphasis on a need to consider implementation through a whole-systems lens those accounts for national level factors (*Tansella & Thornicroft, 2009; Rosen & Salvador-Carulla, 2022*). This is further emphasised when noting the importance of leadership as a core facilitator to successful implementation of such projects as it is necessary to consider the challenge of relying heavily on such factors for the success of a services. Previous literature has noted that the failure to sustain alternative service models can be because of a reliance on charismatic leaders and local champions, without whom they may not thrive (*Lloyd-Evans, Slade, Jagielska & Johnson, 2009)*.

## 4.1. Strengths and Limitations

While the results of this study outline the successful implementation of a range of crisis care models, there are limitations to these findings. Firstly, these results are based on interviews with service managers who provide a single stakeholder perspective, and have been invested in the service, thus are more likely to provide a positive perspective regarding its implementation. Triangulation with quantitative outcome data would be necessary to provide a fuller picture of the impact that such implementation of service has for the wider crisis care pathway and service users receiving this care. While participants were requested to provide further evaluation data to support outcomes and impact monitoring, the resulting return was limited and inconsistent thus could not be utilised for further analysis. Furthermore, as part of a wider evaluation of the services there may be response bias as participants are keen to provide views that outline the success of the project. The study had a low response rate due to a number of projects not partaking for a range of reasons. These services may have had important insights into wider challenges and barriers that the included projects did not experience, thus there is a need to encapsulate these voices in further work into crisis care implementation. Lastly, it is important to highlight that these service implementations occurred within the specific context of the UK and mostly within NHS services which limits the generalisability of such findings to other countries in which healthcare is delivered differently.

A strength is the collaborative, iterative approach to our analysis. Initial deductive coding was conducted by seven of the study researchers. The second stage of inductive coding was conducted by study researchers working together with four Lived Experience Researcher colleagues. The developing coding framework was refined further with input from the wider study team, including colleagues with a variety of mental health clinical experience. This collaboration came with challenges to ensure consistent coding, but the resulting analysis was informed by a range of views and stakeholder perspectives.

### 4.2. Implications for policy, practice and research

Overall, these findings point to the positives associated with adopting innovative approaches through developing new models and services or through the improvement of existing spaces that can help inform future investment and directions for whole systems approaches to both local and national crisis care offers. These findings outline that capital funding projects like this crisis care scheme can stimulate bottom-up initiatives to meet local need, with even relatively modest funding being seen to have the potential to produce benefits to local crisis care systems perceived as meaningful. The findings from this study outline impressive successes across a range of projects that amount from modest funding amounts; from the total investment of £15 million for these BPOS projects, the majority of services kickstarted and have sustained with investment of less that £500,000 (*DHSC, 2017*). Considering the potential impacts that such services may have on service users, local communities and services, such modest investments could support the development of considerable innovation and create alternative pathways, which is highly salient against a backdrop of workforce challenges and economic cuts.

Given the importance of co-production highlighted within our results, future research should consider widening the participation in such evaluations of crisis care to account for service user views and experiences of using these types of services, as well as ensuring that service users are actively involved in the development of such services. Further work is needed to actively utilise these service user voices, as well as quantitative, longitudinal, and health economic approaches to build the evidence base through measuring real-life outcomes and the impact that these improvements and innovations have on the wider crisis care systems to help inform future initiatives and investment.

In addition, this study noted the impact of staff turnover on successful implementation of change in services. Given the growing workforce and resourcing challenges facing mental health services following the COVID-19 pandemic, it is likely that longer term services will experience further staff turnover which will impact motivation and leadership of these innovative models, as well as growing barriers from the ‘outer setting’. As a result, future research should consider longitudinal approaches to examine the longer-term impacts of implementation strategies.

## 5. Conclusion

Against the backdrop of increasing workforce challenges and increasing demand on mental health inpatient beds, as well as the poor experiences reported by service users, there is a recognised need to improve, update, and innovate current crisis care offers. Results from this study suggest that a range of models can help address the heterogenous needs of the local population. Such services can be successfully implemented where they utilise a whole-systems approach, involving service users and relevant professional stakeholders beyond mental health services in planning and developing the service. Flexible, integrated, and collaborative approaches that are informed from the bottom-up are desirable, supported by consistent, active support from senior managers and commissioners.

## Data Availability

All data produced in the present work are contained in the manuscript

## Declarations

### Ethics approval and consent to participate

All procedures performed in studies involving human participants were in accordance with the ethical standards of the institutional and/or national research committee and with the 1964 Helsinki declaration and its later amendments or comparable ethical standards. This study met Health Research Authority criteria for a service evaluation, for which ethical and HRA approvals were not needed. This was independently confirmed by review of the study protocol by the Research Director of the North London Research Consortium (Noclor). All participants provided informed consent prior to enrolment in the study, including consent for publication of anonymised quotes.

### Consent for publication

Not Applicable

### Availability of Data

The data that support the findings of this study are available on request from the corresponding author. The data are not publicly available due to privacy or ethical restrictions.

### Author Contributions

All authors have read and approved the manuscript. Authors BLE, JC, SJ, AS & MS. created the protocol for the study. UF conducted the recruitment for the study, and UF, RA, NL, NA, RS, MS conducted the interviews. UF, RA & PN coded data, and BLE contributed to the thematic analysis. All authors supported drafting and development of the manuscript.

## Acknowledgements

We would like to acknowledge the Mental Health Policy Research Unit team, Lived Experience Working Group members, and Project Working Group who have contributed and supported this work. Special thanks to Beverly Chipp for her contribution to the paper by providing an independent lived experience commentary.

